# Mapping genomic loci prioritises genes and implicates synaptic biology in schizophrenia

**DOI:** 10.1101/2020.09.12.20192922

**Authors:** The Schizophrenia Working Group of the Psychiatric Genomics Consortium, Stephan Ripke, James TR Walters, Michael C O’Donovan

## Abstract

Schizophrenia is a psychiatric disorder whose pathophysiology is largely unknown. It has a heritability of 60-80%, much of which is attributable to common risk alleles, suggesting genome-wide association studies can inform our understanding of aetiology^1^. Here, in 69,369 people with schizophrenia and 236,642 controls, we report common variant associations at 270 distinct loci. Using fine-mapping and functional genomic data, we prioritise 19 genes based on protein-coding or UTR variation, and 130 genes in total as likely to explain these associations. Fine-mapped candidates were enriched for genes associated with rare disruptive coding variants in people with schizophrenia, including the glutamate receptor subunit *GRIN2A* and transcription factor *SP4*, and were also enriched for genes implicated by such variants in autism and developmental disorder. Associations were concentrated in genes expressed in CNS neurons, both excitatory and inhibitory, but not other tissues or cell types, and implicated fundamental processes related to neuronal function, particularly synaptic organisation, differentiation and transmission. We identify biological processes of pathophysiological relevance to schizophrenia, show convergence of common and rare variant associations in schizophrenia and neurodevelopmental disorders, and provide a rich resource of priority genes and variants to advance mechanistic studies.

## INTRODUCTION

Schizophrenia is a clinically heterogeneous disorder, which typically manifests in late adolescence or early adulthood^1^. It is associated with elevated risk of suicide^2^ and serious physical illnesses^3^, reduced life expectancy, and substantial health and social costs. Treatments are at least partially effective in most people, but many have chronic symptoms, and adverse treatment effects are common^4^. There is a need for novel therapeutic target discovery, a process impeded by our limited understanding of pathophysiology.

Much of the variation in risk between individuals is genetic involving alleles that span the full range of frequencies, including large numbers of common alleles^5^ as well as rare copy number variants (CNVs)^6^ and rare coding variants (RCVs)^7,8^. A recent genome-wide association study (GWAS) reported 176 genomic loci containing common alleles associated with schizophrenia^9^ but the causal variants driving these associations and the biological consequences of these variants are largely unknown. To increase our understanding of the common variant contribution to schizophrenia, we performed the largest GWAS to date and analysed the findings to prioritise variants, genes and biological processes that contribute to pathogenesis.

## RESULTS

### Association Meta-Analysis

The primary GWAS was performed on 90 cohorts including 67,390 cases and 94,015 controls (161,405 individuals; equivalent in power to 73,189 each of cases and controls). This was a trans-ancestry analysis with ~80% of the sample of European ancestry and 20% of East Asian ancestry (Sample Supplementary Note). After identical data processing protocols, we conducted within cohort association analyses followed by meta-analysis of 7,585,078 SNPs with MAF ≥ 1%. We identified 294 independent SNPs (linkage disequilibrium (LD) r2 < 0.1) that exceeded genome-wide significance (p<5×10^-8^) (**Figure 1; Supplementary Table 1**).

**Figure 1:**
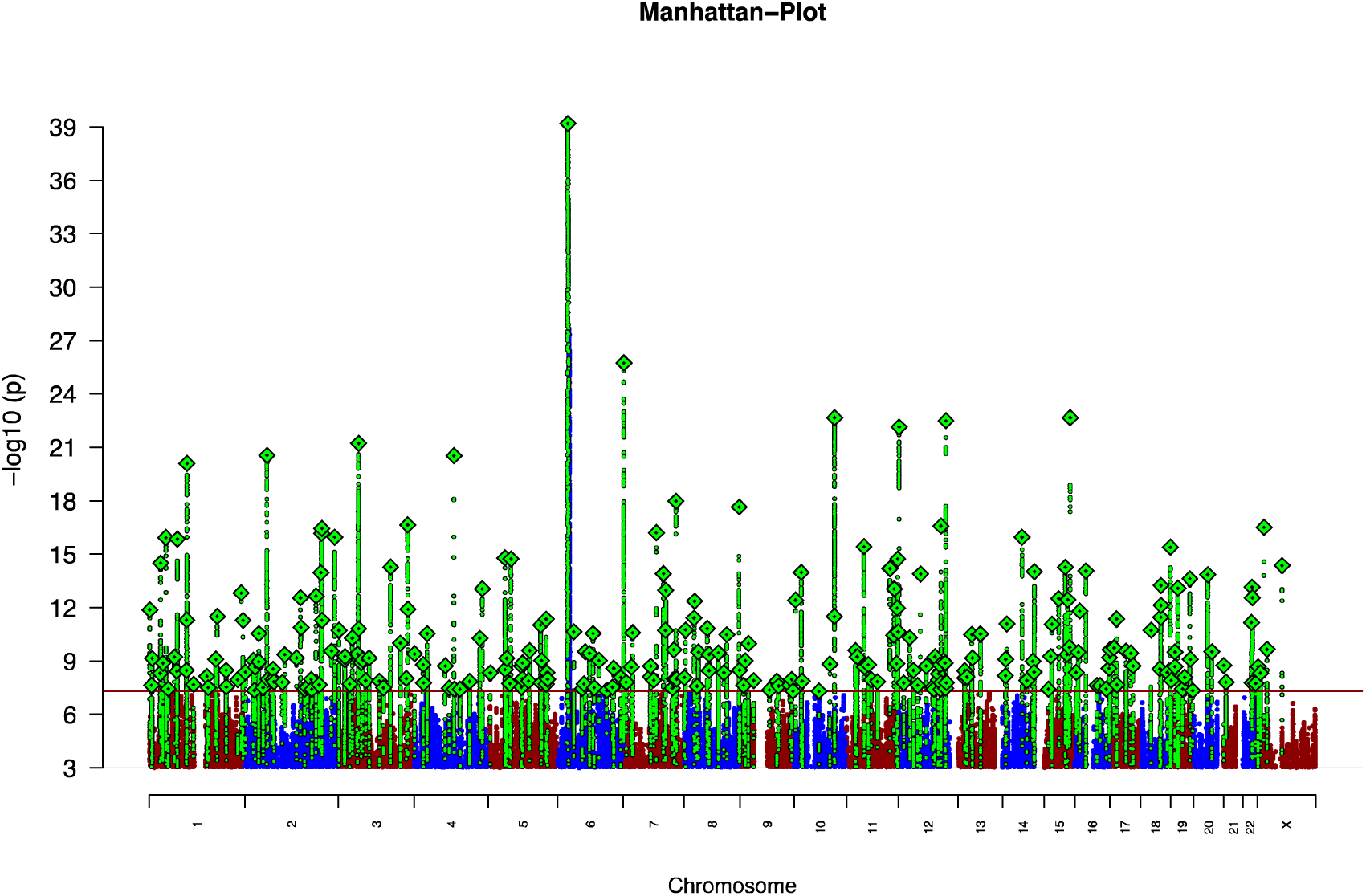
Discovery GWAS Manhattan plot. *The xaxis indicates chromosomal position and the y-axis is the significance of association (–log10(P)). The red line represents genome-wide significance level (5×10*^−^*^8^). SNPs in green are in linkage disequilibrium (LD; R2 >0.1) with index SNPs (diamonds) which represent independent genome-wide significant associations*.

As expected^10^, we observed substantial genome-wide test-statistic inflation above the null (**Extended Data Figure 1**); at least 90% of this is due to polygenicity (**Supplementary Note**) rather than confounders.

**Extended Data Figure 1:**
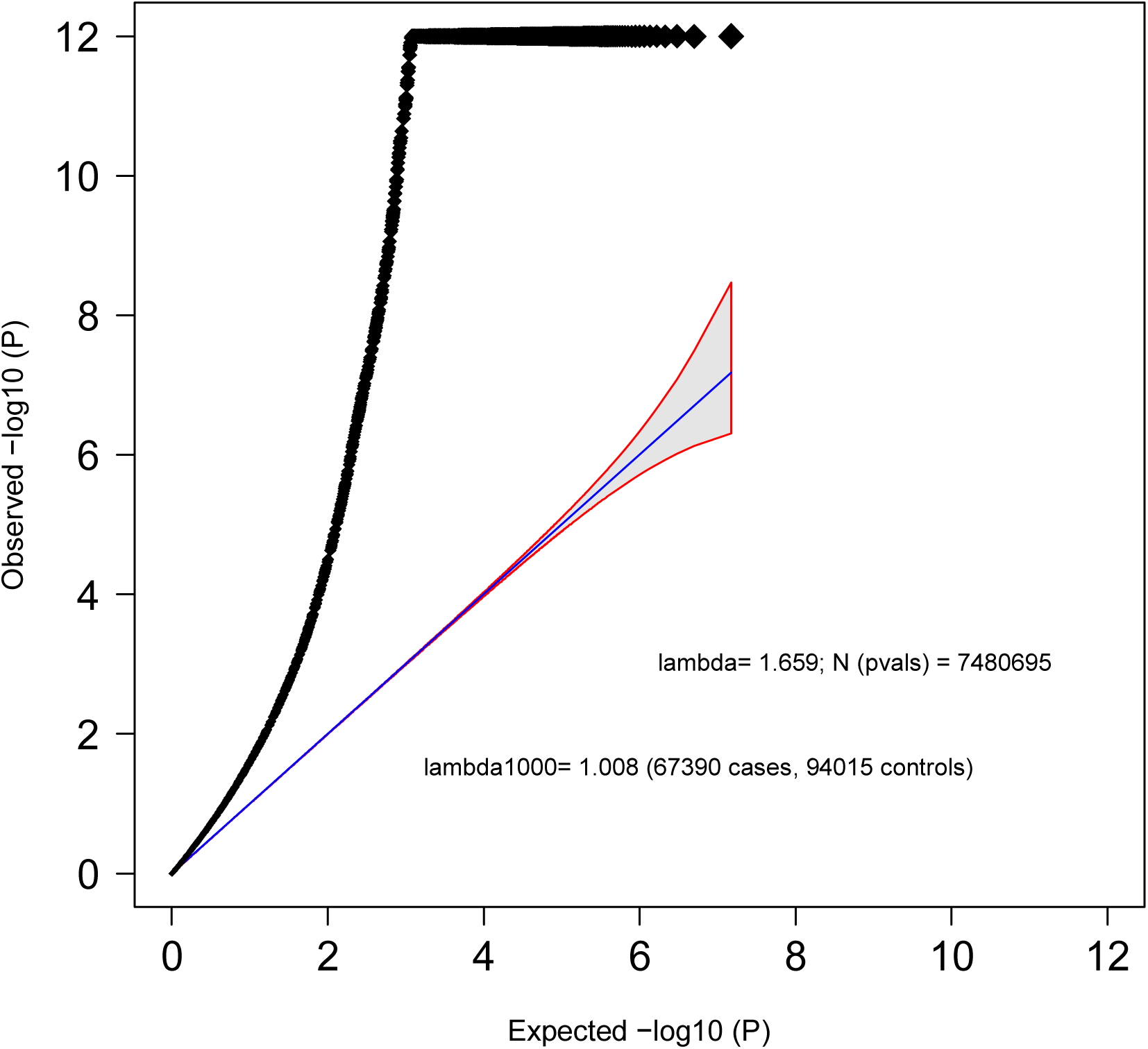
Discovery GWAS Quantilequantile plot. *The xaxis shows the expected –log10(P) values for association under the null distribution given number of independent tests. The y-axis denotes the observed –log10(P). We truncate the Y-axis at –log10(P)=12. The shaded area surrounded by a red line indicates the 95% confidence interval under the null. Lambda is the observed median y χ2 test statistic divided by the median expectedy χ2 test statistic under the null*.

For index SNPs with P<10^-5^, we obtained summary association statistics from deCODE Genetics (1,979 cases, 142,627 controls), the results of which *en masse* were consistent with those from our primary GWAS (**Supplementary Note**). Meta-analysis with deCODE identified 329 LD-independent significant SNPs (**Supplementary Table 2**). These were located in 270 loci (i.e. distinct regions of the genome; **Supplementary Table 3; Supplementary Figures 1-2**). Comparisons with the 128 associations (108 loci) we reported in 2014^11^ are provided in the supplementary note, and the trajectory of associated loci by sample size is depicted in **Extended Data Figure 2;** of note, one previously reported association at rs3768644 (chr2:72.3Mb) is no longer supported^11^.

**Extended Data Figure 2:**
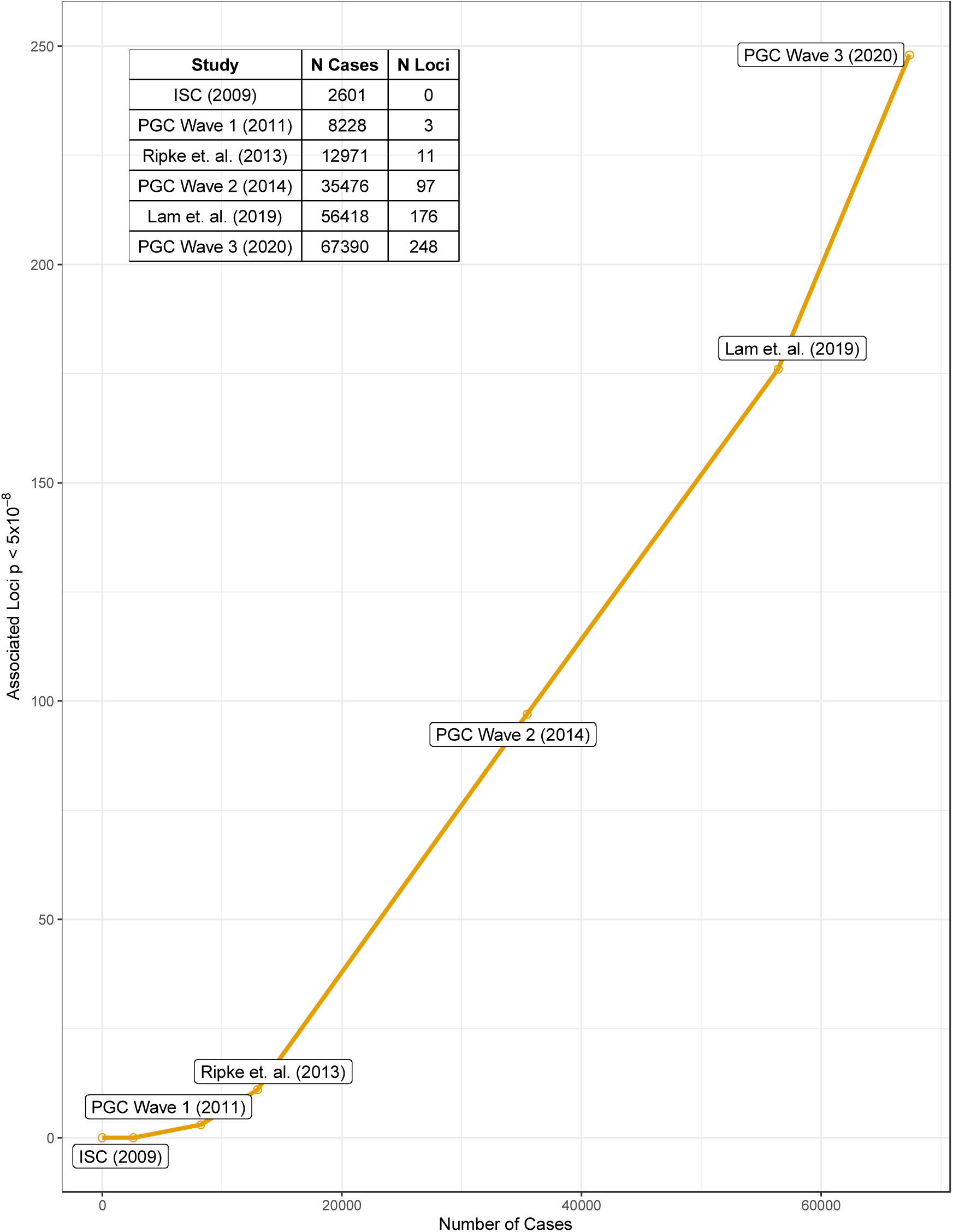
GWAS progress over time. *The relationship of GWAS associations to sample-size is shown in this plot with selected SCZ GWAS meta analyses of the past 11 years. The x-axis shows number of cases. The y-axis shows the number of independent loci discovered with at least one genome-wide significant index SNP in the discovery meta-analysis (e.g. without replication data). The publications listed are ISC (2009)5, PGC Wave 1 (2011)^21^, Ripke et. al. (2013)^52^, PGC Wave 2 (2014)^11^, Lam et. al. (2019)^9^ PGC Wave 3 (2020) - this manuscript. The slope of ~4 newly discovered loci per 1000 cases between 2013 and 2019 increased to a slope of ~6 with the latest sample-size increase*.

The genome-wide significant index SNPs from the primary analyses showed no significant evidence for heterogeneity by sex (**Supplementary Table 4**), and separate GWAS for males and females had a genetic correlation not significantly different from 1 (r_g_=0.992, SE 0.024). These and other analyses (**Supplementary Note**) show that common variant genetic liability to schizophrenia is essentially identical in males and females despite well-established sex differences in age at onset, symptom profile, course, and outcome^12^.

### Heritability and Polygenic Prediction

The proportion of variance in liability attributable to all measured SNPs, the SNP-based heritability (*h*^2^_SNP_), was estimated^13^ to be 0.24 (SE 0.007). Polygenic risk score (PRS) analysis **(Supplementary Note)** explained up to 0.077 of variance in liability (using SNPs with GWAS p-value less than 0.05), and 0.026 when restricted to genome-wide significant SNPs (**Extended Data Figure 5**).

**Extended Data Figure 5:**
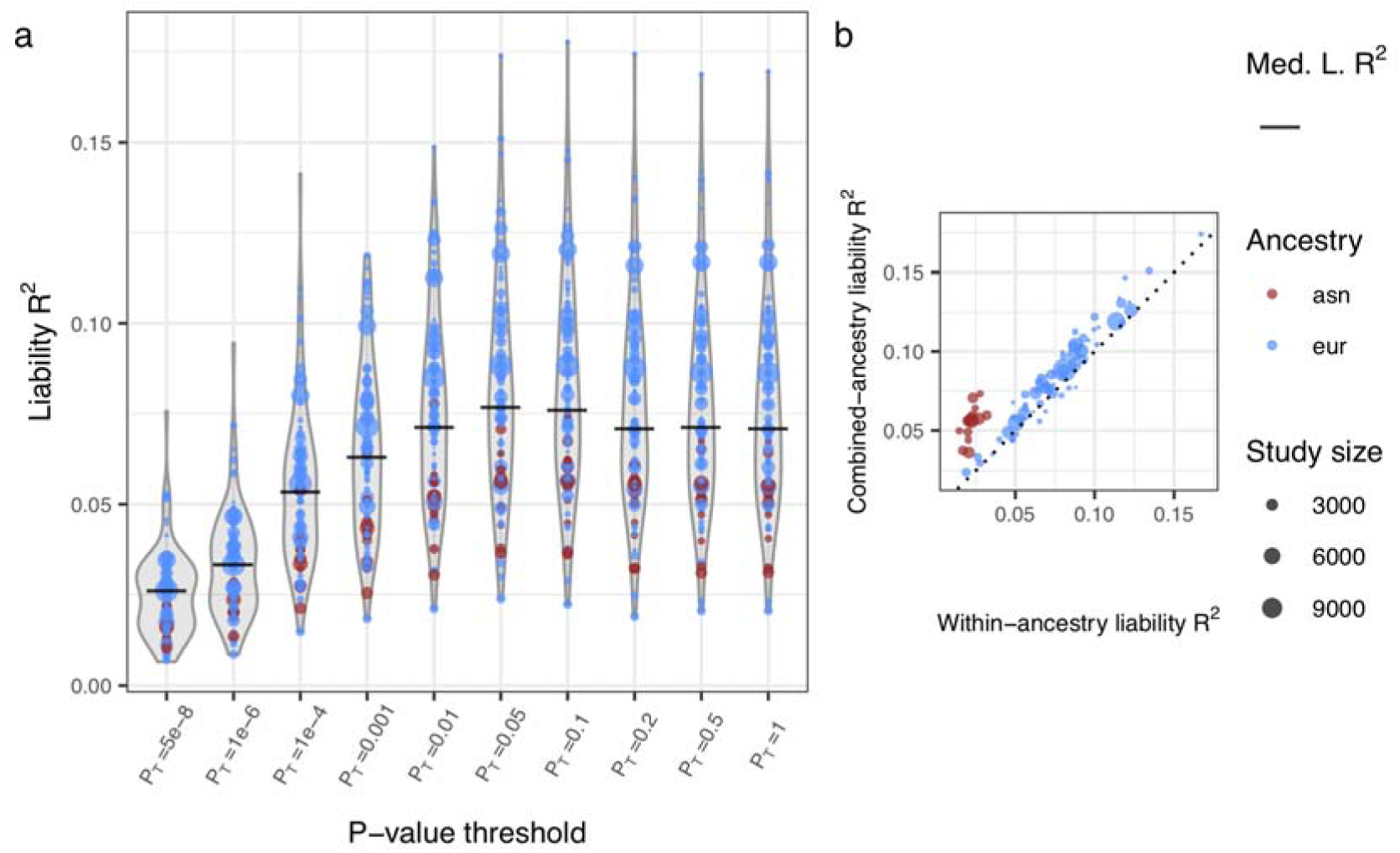
Polygenic risk prediction performance. *A) Distributions of liability R^2^ across 89 left-out-cohorts for polygenic risk scores built from SNPs with different p-value thresholds. Distributions of liability R^2^ (assuming a prevalence of 1%) are shown for each p-value threshold, with point size representing size of the left-out cohort and colour representing ancestry. The sample-size-weighted-mean liability R^2^ is represented as a bar for each method. B) Liability R^2^ of predicted and observed phenotypes in left-out cohorts using threshold p=0.05. The polygenic risk scores that give the x-axis R^2^ are derived from two separate leave-one-out GWAS meta-analyses: one with cohorts of Asian ancestry (with results combined across left-out cohorts), and another with cohorts of European ancestry. The polygenic risk scores that give the y-axis R^2^ are those from A*.

For almost all European and Asian cohorts, PRS had more explanatory power using risk alleles derived from the full combined ancestry GWAS than from the matched ancestry GWAS. PRS explained more variance in liability in cohorts of European ancestry (likely a result of the ancestry composition of the GWAS) but also in samples which by ascertainment are likely to include the most severe cases (i.e. hospitalized patients including those treated with clozapine) **(Supplementary Figure 4, Supplementary Note)**.

A fuller discussion of heritability and polygenic prediction is provided in the **Supplementary Note**; importantly, the liability captured by PRS is insufficient for predicting diagnosis in the general population, with an average Area Under the Receiver Operating Characteristic Curve (AUC) of 0.71. Nevertheless, as a quantitative estimate of liability to schizophrenia, PRS has many applications in research settings, for example for patient stratification, or for identifying correlates of liability in population samples14. In those contexts, PRS indexes substantial differences in liability between individuals; in European samples, compared to the lowest centile of PRS, the highest centile of PRS has an OR for schizophrenia of 44 (95% CI=31-63), and 7.0 (CI 5.8-8.3) when the top centile is compared with the remaining 99% of individuals (**Supplementary Table 5**).

### Mendelian randomisation analyses

Schizophrenia is genetically correlated with several psychiatric, cognitive, and behavioural phenotypes^15^. It is also associated with several non-CNS related traits of clinical importance, including cardiovascular disorders, metabolic syndrome^3,16^, and some autoimmune disorders^17^. The reasons for these associations are unclear, but they may point to traits with causal influences on SZ which could provide clues to prevention. We used 2-sample Mendelian randomisation^18^ using summary-level genomic data for 58 traits to test hypotheses of causal relationships in both directions, that is, where schizophrenia could be a cause or a consequence of the other trait. **(Methods, Supplementary Table 6, Supplementary Note)**.

As reported for complex disorders^16^ we found evidence that alleles with effects on schizophrenia liability frequently increase liability to other traits but that generally these effects are independent of each other (a phenomenon called horizontal pleiotropy) indicated by P_Bias_ in **Extended Data Figure 3**.

**Extended Data Figure 3:**
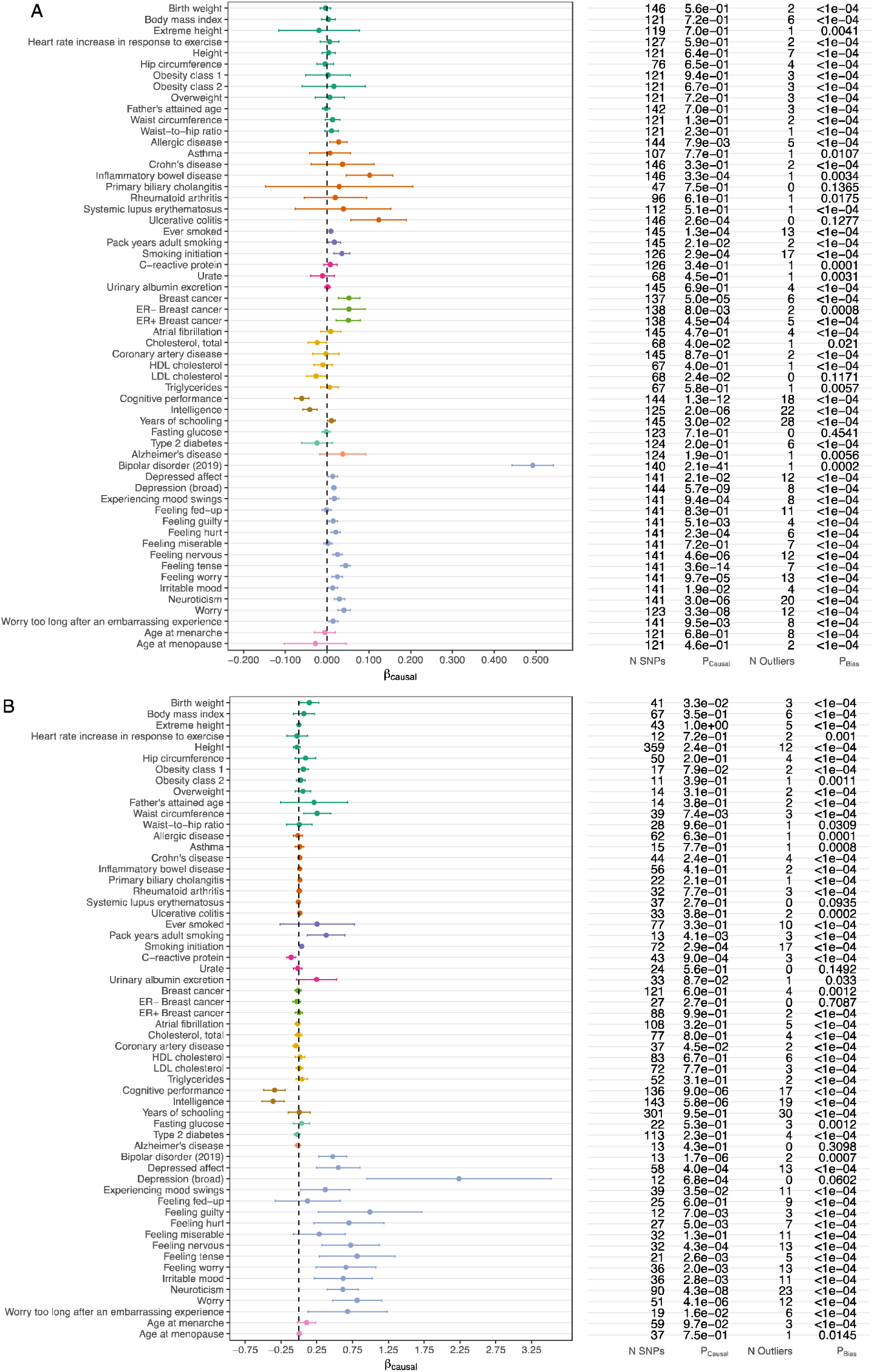
Relationships between schizophrenia and various complex traits and diseases estimated with bidirectional Mendelian randomization. *Panel (a): schizophrenia as the exposure. Panel (b): schizophrenia as the outcome. N SNPs: number of SNPs used as instrumental variables. P_causal_: p-value for causal effects from MR PRESSO^16^ after remo ving SNPs with potential pleiotropic bias. N Outliers: number of SNPs with potential pleiotropic bias identified by MRPRESSO. P_bias_: p-value for MR-PRESSO global test for pleiotropic bias. All tests required a minimum of 10 SNPs for the exposure. All GWAS summary statistics include at least 1,000,000 SNPs and 10,000 samples. Traits of similar types are given the same colour*.

After accounting for pleiotropic bias, 22 of the exposure/outcome tests showed evidence of correlated SNP effect sizes (corrected for 116 tests, P<4.31×10^-4^) (**Extended Data Figure 3**), between schizophrenia and a range of other psychiatric, cognitive or behavioural traits, including bipolar disorder, depression phenotypes, smoking initiation and cognition. However, analyses suggest that correlated genetic effect sizes are not consistent with clear causal relationships between any pair of these traits, and hence are more likely to represent shared aspects of biology (consistent with horizontal pleiotropy). Moreover, as many of these phenotypes also have overlapping features, partial sharing of genetic effects suggests that there may be underlying dimensional traits that are common features of, or increase liability to, these traits rather than one being necessarily causal for another. We did, however, find evidence suggesting schizophrenia increases liability to ulcerative colitis and, as reported before^19^, breast cancer, suggesting the need for studies capable of identifying the potential mechanisms behind these relationships which potentially could be preventable.

### Gene Set Enrichments

To inform hypotheses about aetiopathogenesis we tested for enrichment of associations in sets of genes defined by their expression in tissues, cell types, cell compartments, or annotation to cellular compartments. These analyses test the importance of the characteristics used to define gene sets for disease aetiology, but they do not speak to the involvement or not of individual genes.

### Tissue and cell types

Genes with high relative specificity for bulk expression in many regions of human brain^20^ were strongly enriched for schizophrenia associations **(Extended Data Figure 4)**.

**Extended Data Figure 4:**
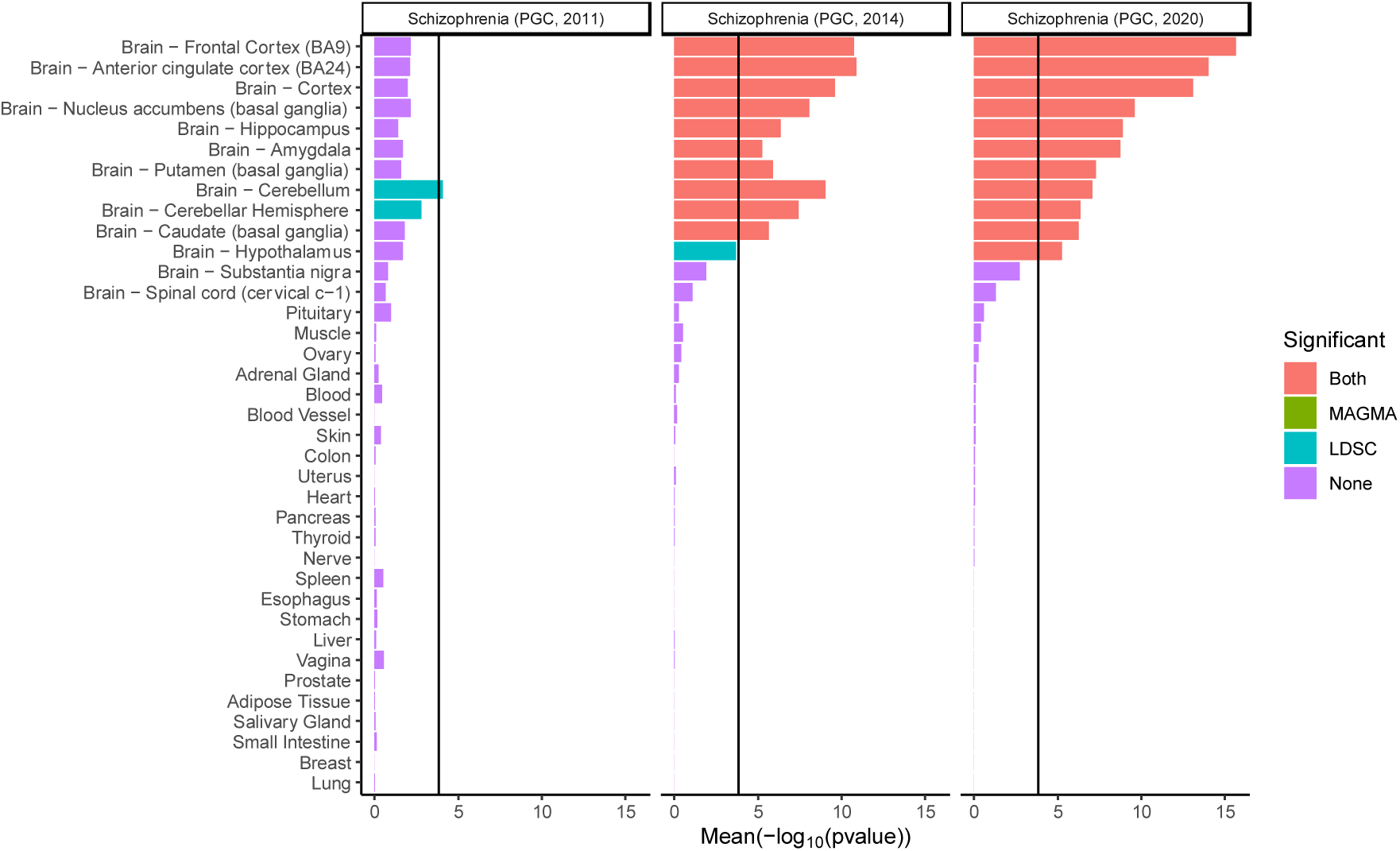
Association between 37 human tissues and schizophrenia. *The mean of strength of association evidence using two enrichment methods (-log_10_P_MAGMA_, -log_10_P_LDSC_) derived from bulk RNA-seq^20^, is shown for each tissue. The bar colour indicates whether the cell type is significantly associated with both methods (i.e. MAGMA and LDSR), one method or none. The black vertical bar represents the significance threshold corrected for the total number of tissues/cell types tested in this study (P = 0.05/341). We also analysed previous waves of PGC schizophrenia GWAS^11,21^ for comparison*.

Comparison with our earlier studies^11,21^ shows increasingly clear contrast between the enrichments in brain and non-brain tissues as sample size increases. Consistent with, but stronger than, previous studies^22^, in single cell expression data^23^, we found associations were enriched in genes with high expression in human cortical inhibitory interneurons and excitatory neurons from cerebral cortex and hippocampus (pyramidal and granule cells) **(Figure 2a)**. From mouse single-cell RNA-seq data^22^, genes highly expressed in excitatory pyramidal neurons from the cortex and hippocampus were strongly associated with schizophrenia **(Figure 2b)** as were those in inhibitory cortical and inhibitory medium spiny neurons, the latter being the predominant cells of the striatum.

**Figure 2:**
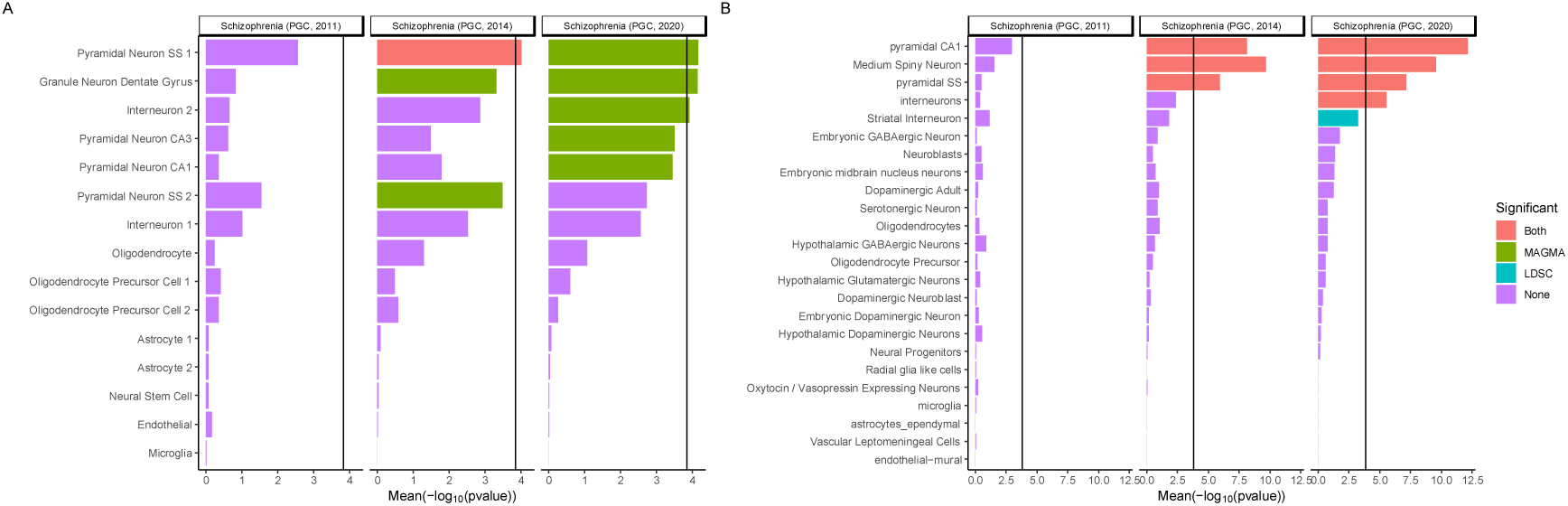
Associations between schizophrenia and cell types from multiple brain regions in human and mouse. *The mean of strength of association evidence using two enrichment methods (-log_10_P_MAGMA_, -log_10_P_LDSC_) between gene expression specificity and schizophrenia is shown (****A****) for 15 human cell types (derived from single nuclei) from the cortex and hippocampus (****B****) and for 24 cell types (derived from single cell RNA-seq) from five different brain regions in mouse (cortex, hippocampus, striatum, midbrain and hypothalamus) and from specific enrichments for oligodendrocytes, serotonergic neurons, dopaminergic neurons and cortical parvalbuminergic interneurons. The bar colour indicates whether the cell type is significantly associated with both methods (MAGMA and LDSC), one method or none. The black vertical bar represents the significance threshold corrected for the total number of tissues/cell types tested in this study (P = 0.05/341). Results obtained for previous iterations of schizophrenia GWAS^11,21^ are shown for comparison. Pyramidal SS: pyramidal neurons from the somato-sensory cortex, Pyramidal CA1: pyramidal neurons from the CA1 region of the hippocampus, Pyramidal CA3: pyramidal neurons from the CA3 region of the hippocampus. Where types of cell (e.g. interneuron) formed sub*220 *clusters in the source data, these are designated by the suffix 1 or 2*.

Across 265 cell types in the mouse central and peripheral nervous system^24^, the strongest enrichments were for genes expressed in glutamatergic neurons located in the deep layers of cortex, amygdala, and hippocampus **(Supplementary Figure 5)**, although enrichments were seen for inhibitory and excitatory neuronal populations more widely (e.g., excitatory neurons from the midbrain, thalamus and hindbrain). Significant enrichments for association signals were not observed for genes with highly specific expression in glia or microglia. Overall, the findings are consistent with the hypothesis that schizophrenia is primarily a disorder of neuronal function, but do not suggest pathology is likely restricted to a circumscribed brain region.

### Biological Processes

Of 7,083 gene ontology (GO) classifications curated to include only annotations with experimental or phylogenetic support, 27 were associated (corrected for multiple testing) with schizophrenia **(Supplementary Table 7)**. Stepwise conditional analysis to minimise redundancy due to genes present in multiple classifications identified 9 associations, all related to neuronal excitability, development, and structure, with prominent enrichment at the synapse. Using the expert-curated synaptic ontology from the SynGO consortium^25^, we found conditionally significant annotations were mainly to postsynaptic terms **(Supplementary Tables 8, 9)**, although enrichments were also found for trans-synaptic signalling and synaptic organisation.

### Fine-mapping

We performed stepwise analyses, conditioning associations in loci on their index SNP (and any subsequent conditionally independent associations) to identify regions that contained independent signals (conditional P<10^-6^) **(Supplementary Note and Supplementary Table 10)**. Where there were independent associations to non-overlapping regions in a locus, we fine-mapped these separately. We used FINEMAP^26^ to assign posterior probabilities (PP) of being causal to SNPs, and constructed credible sets of SNPs that cumulatively capture 95% of the regional PP. We then focussed on loci **(Supplementary Table 3)** that were genome-wide significant in the primary dataset and that were predicted to contain 3 or fewer causal variants **(N=217; Figure 3; Supplementary Tables 11a, 11b)**.

**Figure 3:**
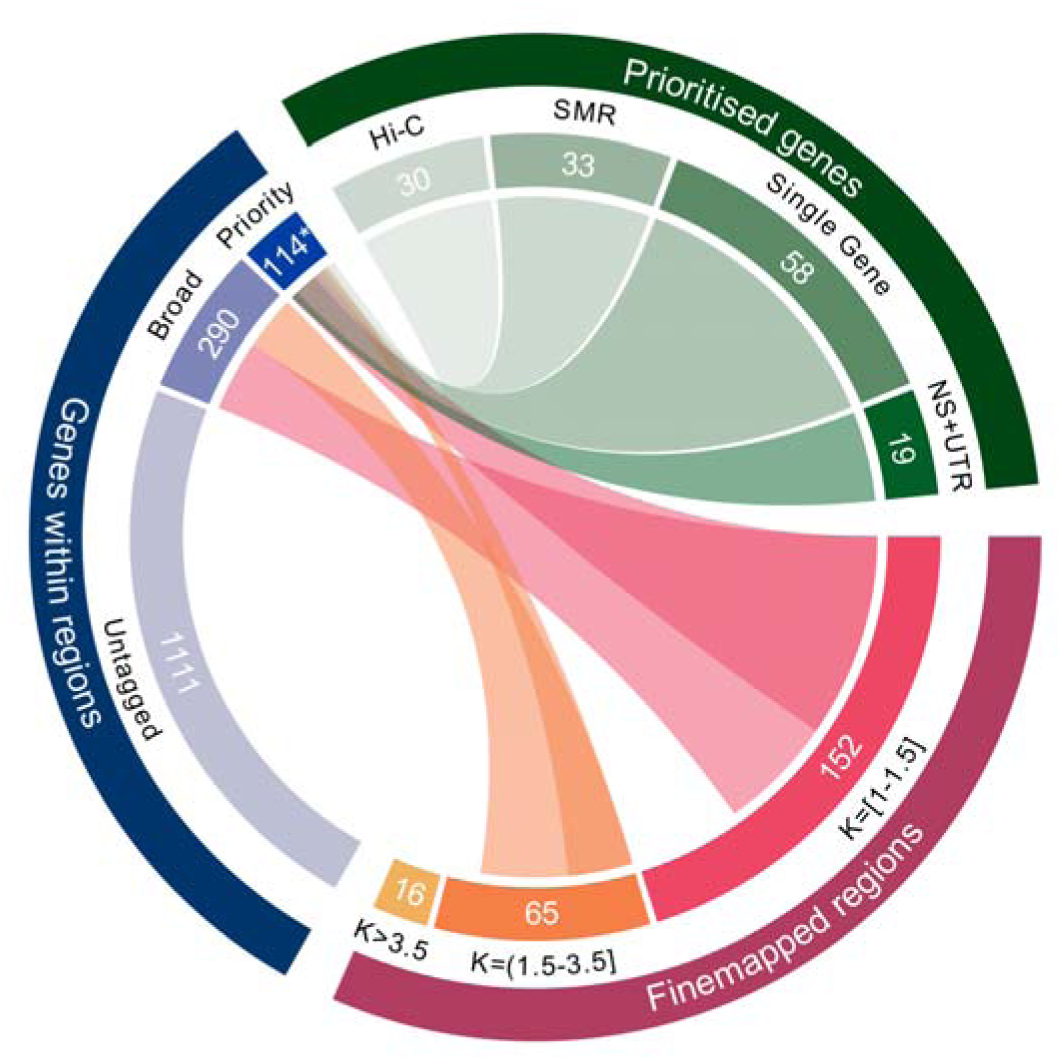
FINEMAP results and overview of the gene prioritisation analysis. *Fine-mapped GWAS regions and number of expected causal variants (K) inferred by FINEMAP. Links are displayed to indicate how the results of FINEMAP were used to select sets of protein-coding genes within associated GWAS loci for further analyses. Two gene sets are shown: “Broad " genes contain at least one FINEMAP credible SNP, while “Priority" genes meet the prioritisation criteria described in the text. The combined total of 114 protein-coding prioritised genes includes 3 that are outside of the 233 fine-mapped regions, but which have expression QTLs within those regions (FOXN2) and Hi-C interactions containing FINEMAP credible SNPs (ZCCHC7, ALG12). Criteria for defining prioritised genes are also shown. “NS+UTR": Genes containing a FINEMAP credible nonsynonymous or UTR SNP with posterior probability greater than 10%. “Single Gene”: Genes in which the entire FINEMAP credible SNP set was contained within one gene. “SMR”: protein coding genes in which SMR analysis points towards a single gene at a locus, or where the credible eQTL causal SNPs capture >50% of the FINEMAP posterior probability. “Hi-C”: protein coding genes with SMR evidence, and where enhancer-promoter or promoter-promoter interaction exists between segments of DNA containing FINEMAP credible SNPs. Note these criteria are not mutually exclusive and thus individual genes might fulfil more than one criterion (****Supplementary Table 20****). For clarity, links to genes within associated GWAS loci that were not tagged by FINEMAP credible SNPs are not depicted*.

For 32 loci, the 95% credible set contained 5 or fewer SNPs (**Supplementary Table 11c)** and for 8 loci, only a single SNP. We draw particular attention to intronic SNP rs4766428 (PP>0.97) at *ATP2A2* which encodes a Sarcoplasmic/Endoplasmic Reticulum Calcium pump. Mutations in *ATP2A2* cause Darier Disease^27^, which co-segregates with bipolar disorder in several multiplex pedigrees, and is also associated with bipolar disorder and schizophrenia at a population level^28^. Given enrichment for associations in and around voltage gated calcium channels **(Supplementary Tables 3 and 7)**, *ATP2A2* may be involved in pathogenesis through its role in regulating cytoplasmic calcium in neurons.

We identified a fine-mapped set comprising genes with at least one credible SNP. We *prioritised* those containing a) at least one nonsynonymous (NS) or untranslated region (UTR) variant with a PP≥0.1 **(Supplementary Table 12)** or (b) the entire credible set was annotated to that gene **(Supplementary Table 13)**. Genes that are expressed in brain, and that are relatively intolerant to loss-of-function mutations, are known to be enriched for schizophrenia associations (^29^ and **Figure 4A**). Genes prioritised by FINEMAP were further enriched for these properties compared to other genes in the loci **(Figure 4B; Supplementary Table 14; Supplementary Note)** supporting our prioritisation strategy.

**Figure 4:**
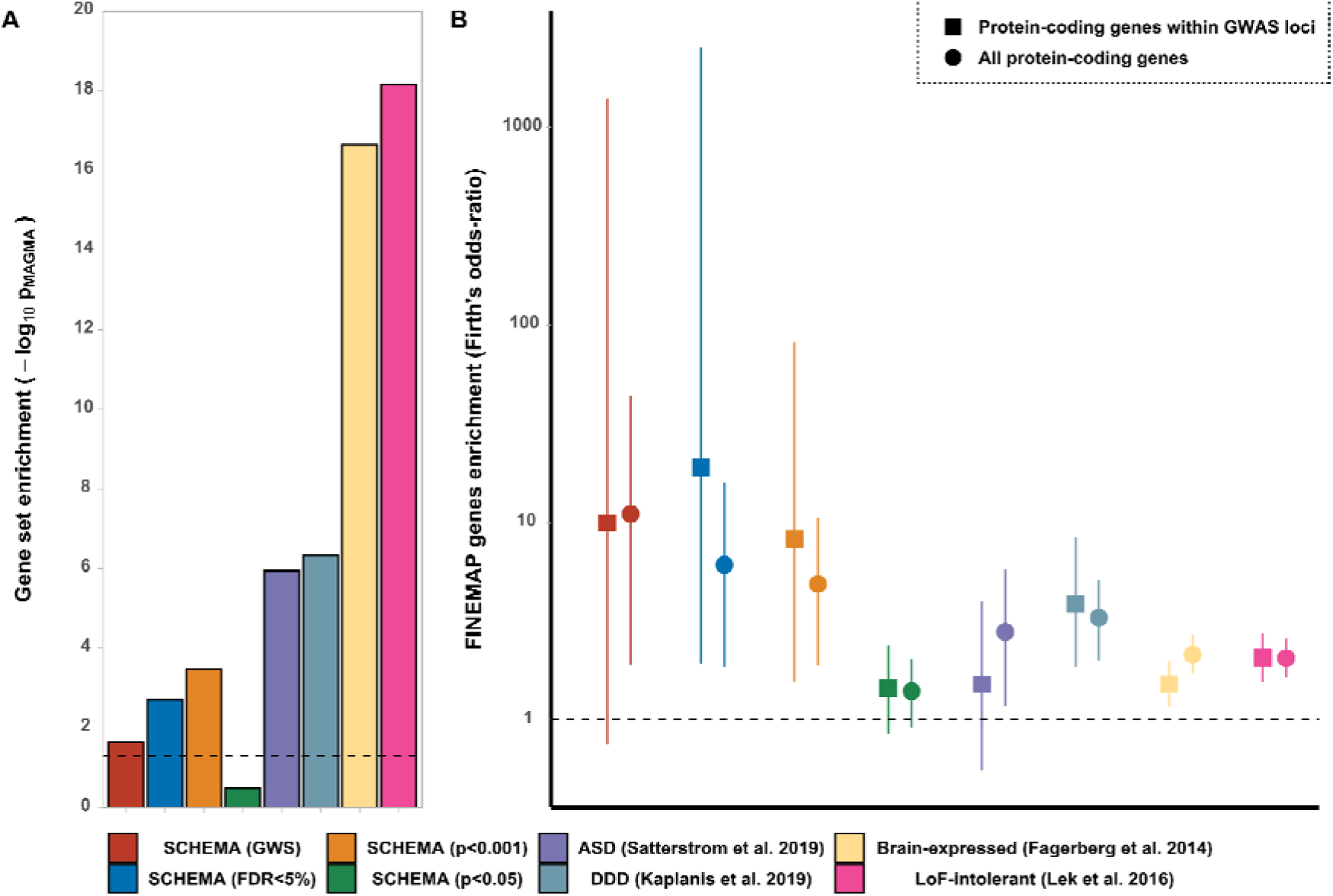
Gene set tests. *Gene sets tested were retrieved from genome and exome-wide sequencing studies of schizophrenia (SCHEMA; companion paper), autism-spectrum disorder^37^ and developmental disorders^36^. For reference, additional sets representing LoF-intolerant genes53 and brain-expressed genes^54^ are also shown. A: MAGMA gene set enrichment analysis, dotted line indicates nominal significance (p=0.05). B: Logistic regression (with Firth’s bias reduction method) showing the odds-ratio (and 95% confidence interval) of the association between genes containing at least 1 credible FINEMAP SNP and genes from the sets indicated. Odds-ratios are relative to the remaining protein-coding genes within GWAS loci (squares) or across the genome (circles). Dotted line indicates no enrichment. Analyses are adjusted for gene size*.

We prioritised 19 genes (all protein coding by definition) based on NS or UTR variants (**Supplementary Table 12**), including *SLC39A8*, which mediates zinc and manganese uptake, in which rs13107325, previously a moderately high credible SNP^29^, is now strongly supported as responsible (PP > 0.99). The associated T allele has pleiotropic associations of potential relevance to schizophrenia^30^. Other non-synonymous variants with high PP were found in genes with minimal functional characterization including *THAP8* (2 alleles in perfect LD), *WSCD2*, and in 2 E3 ubiquitin ligases *PJA1* and *CUL9*. Also notable for reasons discussed above (see *ATP2A2*), a nonsynonymous and a UTR variant prioritised the voltage gated calcium channel subunit *CACNA1I*. A mixture of missense and UTR variants prioritised *interferon regulatory factor 3 (IRF3;* cumulative PP=0.49) which promotesinterferon expression^31^ while *KLF6*, a transcription factor, was highlighted by three variants in the 3’ UTR (cumulative PP > 0.99). Interestingly, IRF deficiency has been functionally and genetically linked to herpes simplex encephalitis^32^, which can present with schizophrenia-like features. Finally, we identified 65 genes (58 protein coding) in which the 95% credible set is restricted to a single gene (Supplementary Table 13).

### Common and Rare Variant Associations with Schizophrenia

The Schizophrenia Exome Sequencing Meta-Analysis (SCHEMA) consortium (companion paper) identified 32 genes with damaging ultra-rare mutations associated with schizophrenia (FDR<0.05), including 10 at exome-wide significance. Using MAGMA^33^, we found both sets of genes were enriched for common variant associations, as were more weakly associated SCHEMA genes down to uncorrected P<0.001 (**Figure 4A, Supplementary Tables 15, 16**). The fine-mapped set was also enriched for SCHEMA genes relative to other genes at associated loci (**Figure 4B; Supplementary Table 16**). Given established rare variant overlaps between schizophrenia, autism spectrum disorder (ASD) and developmental disorder (DD) ^8,34,35^, we tested for and found the fine-mapped set was also enriched for genes in which rare variants increase risk of these disorders^36,37^ (**Figure 4, Supplementary Tables 15, 16**). These convergences suggest rare variant data can inform gene prioritisation at GWAS loci.

Of the 10 exome-wide significant genes identified by SCHEMA, two were prioritised candidates from fine-mapping; *GRIN2A* encoding a glutamatergic NMDA receptor subunit, and *SP4*, a transcription factor highly expressed in brain and which is regulated by NMDA transmission, and also regulates, NMDA receptor abundance^38^. Two other genes implicated by SCHEMA at FDR<0.05 had support from fine-mapping: *STAG1* which is involved in controlling chromosome segregation and regulating expression, and *FAM120A*, which encodes an RNA binding protein. SNPs mapping to these genes captured 85% and 70% of the FINEMAP PP respectively (**Supplementary Table 11b**). The prioritised fine-mapped set also contained 8 genes implicated in ASD and/or DD; 5 transcriptional regulators *(FOXP1, MYT1L, RERE, BCL11B, KANSL1)*, the well-known candidate *CACNA1C^52^*, and genes mentioned elsewhere in this paper (*GRIN2A* and *SLC39A8*).

### Prioritisation by Gene Expression

It is thought that common variant associations in schizophrenia are frequently mediated by eQTLs, that is, variants that influence gene expression^39^. We used summary-based Mendelian randomisation (SMR)^40^ to detect GWAS associations that co-localise with eQTLs (from adult brain^41^, fetal brain^42^ or whole blood^43^) and the HEIDI test^40^ to reject co-localisations due to LD between distinct schizophrenia-associated and eQTL variants **(Supplementary Table 17)**. To retain brain relevance, we considered only findings from blood that replicated in brain. After removing duplicates identified in multiple tissues **(Supplementary 18a-c)**, we identified 116 SMR-implicated genes **(Supplementary Table 18d);** the use of alternative methodologies supported the robustness of the SMR findings **(Supplementary Note and Supplementary Table 18e)**.

We used three approaches to prioritise genes from these 116 candidates **(Supplementary Note)**. We identified (i) 33 genes as the single SMR-implicated gene at the locus or through conditional analysis of a locus containing multiple candidates **(Supplementary Table 18f; Supplementary Note)**: (ii) 18 genes where the putatively causal eQTLs captured 50% or more of the FINEMAP posterior probability (**Supplementary Table 18g**): (iii) 30 genes where chromatin conformation analysis (Hi-C analysis of adult and fetal brain) suggested a promoter of that gene interacted with a putative regulatory element containing a FINEMAP credible SNP **(Supplementary Table 19)**^44^.

After removing duplicates, there were 60 SMR-prioritised genes **(Supplementary Table 20)**. The neurodevelopmental disorder gene *RERE* (over-expression in schizophrenia) was prioritised by all 3 strategies and separately by fine-mapping (see above). Others with strong evidence **(Supplementary Table 18g)** include *ACE* encoding angiotensin converting enzyme, the target of a major class of anti-hypertensive drugs (schizophrenia underexpression), *DCLK3* encoding a neuroprotective kinase^45^(schizophrenia under-expression) and *SNAP91* (discussed below; schizophrenia over-expression).

Combining all approaches, FINEMAP and SMR, we prioritised 130 genes of which 114 are protein coding (**Figure 3; Extended Data Table 1**).

### Synaptic Location and Function of Prioritised Genes

Given the implication of synaptic pathology from the enrichment tests, we examined prioritised genes in the context of synaptic location and function in the SynGO database^25^. Of the 114 proteins encoded, 13 have synaptic annotations **(Supplementary Table 21)**; 6 are postsynaptic, 5 are both pre- and post-synaptic, 1 is presynaptic, and 1 gene (MAPK3) is not mapped to any specific compartment (**Figure 5B**).

**Figure 5:**
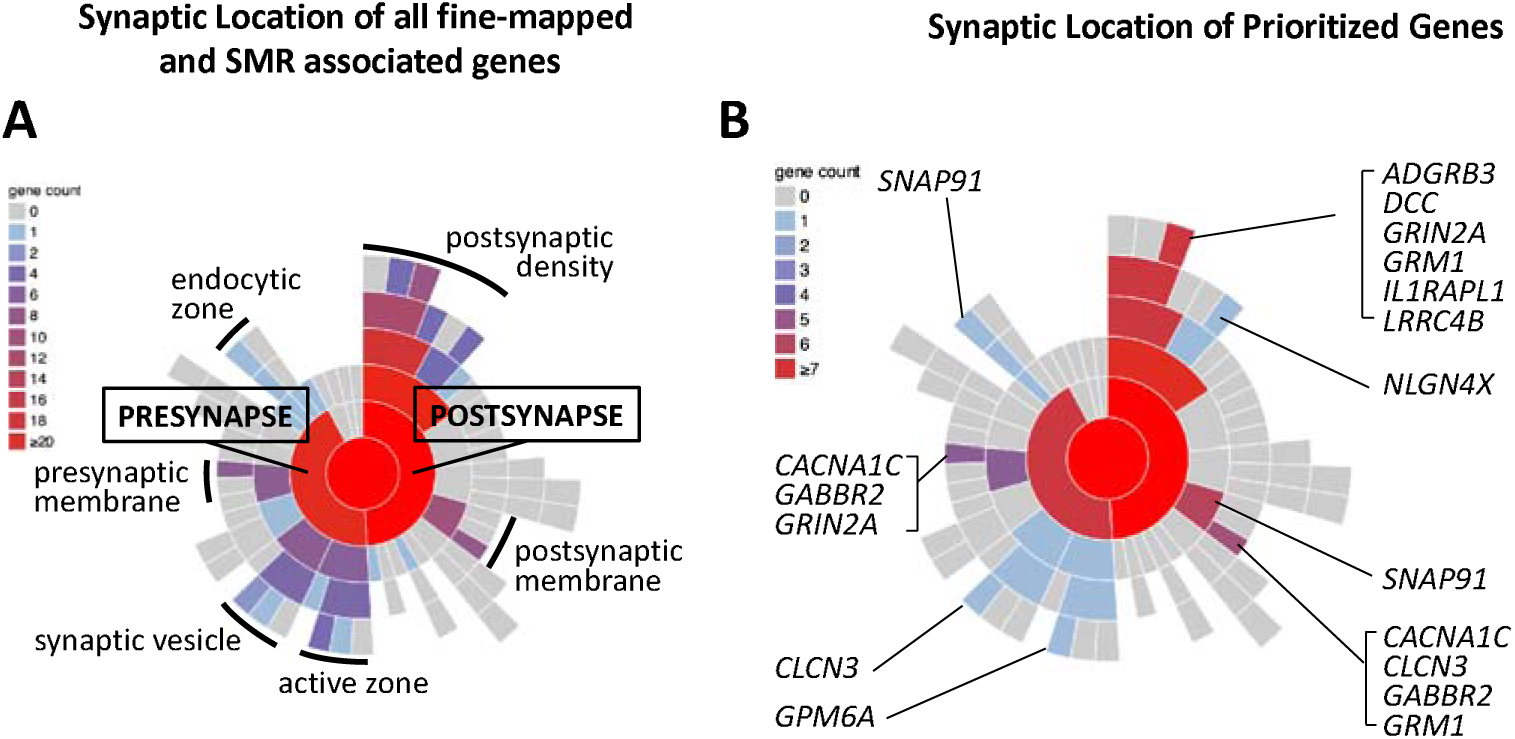
Mapping of all FINEMAP/SMR genes (A) and prioritized genes (B) to synaptic locations using SYNGO. *Sunburst plots depict synaptic locations with child terms in concentric rings, starting with the synapse (center), pre- and postsynaptic locations in the first ring and child terms in subsequent ring. The number of genes in each term is indicated by the colour scheme in the legend. FINEMAP/SMR genes are those tagged by at least one credible SNP identified by FINEMAP or which are associated using SMR (N=437protein coding genes) of which N=50 are SynGO annotated, 44 to cellular components. Prioritized genes are the subset of protein coding genes identified as likely to be causal (**Extended Data Table 1**; N=114) using the prioritization criteria described in the main text of which 13 are SynGO annotated, 12 to cellular components. Genes were annotated using the SYNGO synaptic location database (www.svngoportal.ore) which is exclusively based on expert annotation of published evidence (see Koopmans et al^25^)*.

The results are consistent with the genome-wide enrichment tests pointing to postsynaptic pathology, but the fact that many prioritised genes have additional locations suggests presynaptic pathology may also be involved. The encoded proteins map to 17 unique biological terms in the hierarchy **(Supplementary Table 21**), but there are specific themes. Multiple genes encode receptors and ion channels, including voltage-gated calcium and chloride channels *(CACNA1C, CLCN3)*, metabotropic receptors (glutamate and GABA), and the ligand-gated NMDA receptor subunit *(GR1N2A)*. Others involve proteins playing a role in synaptic organisation and differentiation *(ADGRB3, LRRC4B, GPM6A, 1L1RAPL1)*, the trans-synaptic signalling complex *(1L1RAPL1, SNAP91)* and modulation of chemical transmission *(MAPK3, DCC, CLCN3). ADGRB3*, encoding adhesion G protein-coupled receptor B3, is a receptor for ligands of the secreted complement factor family and together they control the synaptic connectivity and refinement^46,47^. This is notable given that the complement component 4 genes, which have been shown to account for some of the association signal across the MHC locus^49^ and are part of a classical complement pathway in which multiple complement proteins regulate synaptic refinement^50^. The diversity of synaptic proteins identified in this study suggests multiple functional interactions of schizophrenia risk converging on synapses. It remains to be determined whether these proteins co-act together at specific synapse types, or whether the diversity points to multiple types in different brain regions.

## DISCUSSION

We have performed the largest GWAS of schizophrenia to date and in doing so, identify a substantial increase in the number of associated loci. We show that genes we prioritise within associated loci by fine-mapping are highly enriched for those with an increased burden of rare deleterious mutations in schizophrenia, and identify *GRIN2A*, *SP4*, *STAG1*, and *FAM120A* as specific genes where the convergence of rare and common variant associations strongly supports their pathogenic relevance for the disorder. Importantly, this convergence also implies that the pathogenic relevance of altered function of these genes extends beyond the small proportion of cases who carry rare mutations in them. We also demonstrate that common variant schizophrenia associations, and more importantly, 8 prioritised associated genes that likely explain associations, overlap with genes implicated by rare coding variants in neurodevelopmental disorders, opening the door for exploiting the increasing power of rare variant studies of those disorders to further prioritise genes from GWAS studies.

The power of the present study allowed us to show enrichment of common variant associations restricted to genes expressed in CNS neurons, both excitatory and inhibitory, and fundamental biological processes related to neuronal function. This points to neurons as the most important source of pathology in the disorder. We also show that genes with high relative specificity for expression in almost all tested brain regions are enriched for genetic association. This suggests that abnormal neuronal function in schizophrenia is not confined to a small number of brain structures, which in turn might explain its diverse psychopathology, its association with a broad range of cognitive impairments, and the lack of regional specificity in neuroimaging measures associated with the disorder^1^.

Schizophrenia pathophysiology is likely to extend beyond the synapse. Nevertheless, the concentration of associations we show in genes with pre- and post-synaptic locations (including glutamate and GABA receptors and their associated proteins) and with functions related to synaptic organisation, differentiation and transmission, point to these neuronal compartments and their attendant functions as being of central importance. This is further supported by studies showing that CNVs^51^ and rare damaging coding variants in genes with similar functions, and even some of the same genes (SCHEMA; companion paper) substantially increase risk of schizophrenia. Genomic studies of all designs now converge in suggesting research aiming for a mechanistic understanding of the disorder should be targeted towards these areas of biology; the large number of candidates identified here offers an unprecedented empirically-supported resource for that endeavour.

### Ethics

The study protocols were approved by the institutional review board at each centre involved with recruitment. Informed consent and permission to share the data were obtained from all subjects, in compliance with the guidelines specified by the recruiting centres’ institutional review boards. Genotyping of samples recruited in mainland China were processed and analysed by Chinese groups on Chinese local servers, to comply with the Human Genetic Resources Administrative Regulations. Only summary statistics, with no individual-level data, were included in the final study from samples recruited from mainland China.

## Data Availability

Clumped and genome-wide summary statistics are available at the PGC UNC link provided below.

https://www.med.unc.edu/pgc/download-results/

## Acknowledgements

The National Institute of Mental Health (USA) provides core funding for the Psychiatric Genomics Consortium was Award Number U01MH109514. The content is the responsibility of the authors and does not necessarily represent the official views of the National Institutes of Health. We acknowledge Pamela Sklar (deceased); as one of the PIs of that award, her contribution was substantial. The work of the contributing groups was supported by numerous grants from governmental and charitable bodies as well as philanthropic donation (details to be provided). We also acknowledge Ed Scolnick, Chief Scientist Emeritus, Stanley Center of the Broad Institute, whose support for this study was vital to its success. We also acknowledge the Wellcome Trust Case Control Consortium for the provision of control genotype information. Membership of the Psychosis Endophenotype International Consortium, the SynGO consortium, the PsychENCODE Consortium, the eQTLGen consortium, the BIOS Consortium and the Indonesia Consortium are provided in the accompanying author and consortium XL file.

S.X. also gratefully acknowledges the support of the National Natural Science Foundation of China (NSFC) grant (31525014, 91731303, 31771388, 31961130380, and 32041008), the UK Royal Society-Newton Advanced Fellowship (NAF\R1\191094), Key Research Program of Frontier Sciences (QYZDJ-SSW-SYS009) and the Strategic Priority Research Program (XDB38000000) of the Chinese Academy of Sciences, and the Shanghai Municipal Science and Technology Major Project (2017SHZDZX01).

This study is supported by Research Council of Norway (283798, 262656, 248980, 273291, 248828, 248778, 223273); KG Jebsen Stiftelsen, South-East Norway Health Authority, EU H2020 # 847776.

Dr. Faraone is supported by the European Union’s Seventh Framework Programme for research, technological development and demonstration under grant agreement no 602805, the European Union’s Horizon 2020 research and innovation programme under grant agreements No 667302 & 728018 and NIMH grants 5R01MH101519 and U01 MH109536-01.

This study was supported by FAPESP - Fundação de Amparo à Pesquisa do Estado de Sao Paulo (Brazil) - Grant numbers: 2010/08968-6 (S.I.B.); 2014/07280-1 (S.I.B.); 2007/58736-1 (M AC S); 2011/50740-5 (R.A.B.); 2016/04983-7 (J.J.M.); 10/19176-3 (V.K.O. & S.I.B.); 12/12686-1 (M.L.S. & S.I.B.); CAPES - Coordenafao de Aperfeiçoamento de Pessoal de Nível Superior Code 001.

The Singapore team (Lee Jimmy, Liu Jianjun, Sim Kang, Chong Siow Chong, Mythily Subramanian) acknowledges the National Medical Research Council Translational and Clinical Research Flagship Programme (grant number: NMRC/TCR/003/2008).

Supported by LM2018132, CZ.02.1.01/0.0/0.0/18_046/0015515 and IP6003 - VZFNM00064203 to MM Jr.

Dr. Arango has been funded by the Spanish Ministry of Science and Innovation. Instituto de Salud Carlos III (SAM16PE07CP1, PI16/02012, PI19/024), co-financed by ERDF Funds from the European Commission, “A way of making Europe”, CIBERSAM. Madrid Regional Government (B2017/BMD-3740 AGES-CM-2), European Union Structural Funds. European Union Seventh Framework Program; and European Union H2020 Program under the Innovative Medicines Initiative 2 Joint Undertaking (grant agreement No 115916, Project PRISM, and grant agreement No 777394, Project AIMS-2-TRIALS), Fundación Familia Alonso and Fundación Alicia Koplowitz.

E. Bramon was supported by the Medical Research Council (G0901310 and G1100583), the Wellcome Trust (085475/B/08/Z, 085475/Z/08/Z), Mental Health Research UK, British Medical Association’s Margaret Temple Fellowship, and the NIHR Biomedical Research Centre at University College London Hospitals NHS Foundation Trust and University College London.

M.Dolores Moltó is funded by the European Regional Development Fund (ERDF)-Valencian Community 2014-2020, Spain.

## METHODS

### Overview of Samples

Details of each of the samples (including sample size, ancestry, and whether included in the previous publication by the PGC) are given as a **Sample Supplementary Note**. From 90 cohorts, we performed GWAS on 161,405 unrelated subjects; 67,390 schizophrenia/schizoaffective disorder cases and 94,015 controls. A parent-proband trio is considered to comprise one case and one control. Approximately half (31,914 cases and 47,176 controls) of the samples were not included in the previous GWAS of the PGC^1^. Around 80% of the cases were of European Ancestry, and the remainder were of East Asian ancestry^2^. Variants showing evidence for association (P< 1×10^-5^) were further meta-analysed with an additional dataset of 1,979 cases and 142,627 controls of European ancestry obtained from deCODE genetics, thus the final analysis represents 306,011 diploid genomes.

### Association Analysis

#### Quality Control

Quality control was performed on the cohorts separately according to standards developed by the Psychiatric Genomics Consortium (PGC)^3^ including SNP missingness < 0.05 (before sample removal); subject missingness < 0.02; autosomal heterozygosity deviation (| F_het_ | < 0.2); SNP missingness < 0.02 (after sample removal); difference in SNP missingness between cases and controls < 0.02; and SNP Hardy-Weinberg equilibrium (HWE: *P* > 10^−6^ in controls or *P* > 10^−10^ in cases). For family-based cohorts we excluded individuals with more than 10,000 Mendelian errors and SNPs with more than 4 Mendelian errors. For X-Chromosomal genotypes we applied an additional round of the above QC to the male and female subgroups separately.

#### Imputation

Genotype imputation of case-control cohorts was performed using the pre-phasing/imputation stepwise approach implemented in EAGLE 2^4^ / MINIMAC3^5^ (with 132 genomic windows of variable size and default parameters). The imputation reference consisted of 54,330 phased haplotypes with 36,678,882 variants from the publicly available HRC reference, release 1.1 (URL: https://ega-archive.org/datasets/EGAD00001002729). Chromosome X imputation was conducted using individuals passing quality control for the autosomal analysis. ChrX imputation and association analysis was performed separately for males and females. For trio-based cohorts, families with multiple (N) affected offspring were split into N parent-offspring trios, duplicating the parental genotype information. Trios were phased with SHAPEIT 3^6^. We created pseudo-controls based on the non-transmitted alleles from the parents. Phased case-pseudo-control genotypes were then taken forward to the IMPUTE4 algorithm^7^ into the above HRC reference panel.

#### Principal Component Analysis (PCA) and Relatedness Checking

We performed PCA for all 90 cohorts separately using SNPs with high imputation quality (INFO >0.8), low missingness (<1%), MAF>0.05 and in relative linkage equilibrium (LD) after 2 iterations of LD pruning (r2 < 0.2, 200 SNP windows). We removed well known long-range-LD areas (MHC and chr8 inversion). Thus, we retained between 57K and 95K autosomal SNPs in each cohort. SNPs present in all 90 cohorts (N=7,561) were used for robust relatedness testing using PLINK v1.9^8^; pairs of subjects with PIHAT > 0.2 were identified and one member of each pair removed at random, preferentially retaining cases and trio members over case-control members.

#### Association / Meta-analysis

In each individual cohort, association testing was based on an additive logistic regression model using PLINK^8^. As covariates we used a subset of the first 20 principal components (PCA), derived within each cohort. By default, we included the first 4 PCAs and thereafter every PCA that was nominally significantly associated (p<0.05) to case-control status. PCAs in trios were only used to remove ancestry outliers. We conducted a meta-analysis of the results using a standard error inverse-weighted fixed effects model. For chrX, gene dosages in males were scored 0 or 2, in females, 0/1/2. We summarised the associations as number of independently associated index SNPs. Index SNPs were LD independent and had r2 < 0.1 within 3 Mb windows. We recorded the left and rightmost variant with r2<0.1 to an index SNP to define an associated clump. After adding a 50kb window on each side of the LD clump we merged overlapping LD-clumps to a total of 248 distinct genomic loci (5 on the X-chromosome) with at least one genome-wide significant signal.

Due to the strong signal and high linkage disequilibrium in the MHC, only one SNP was kept from the extended MHC region (chr6:25-35Mb).

We additionally examined the X chromosome for evidence of heterogeneity between the sexes and X chromosome dosage compensation using the methods described by Lee and colleagues ^9,10^ (**Supplementary Note**). To minimise possible confounding effects of ancestry on effect sizes by sex, we restricted this analysis to those of European ancestry.

We obtained summary association results from deCODE genetics for 1,187 index SNPs (P < 1×10^-5^) based on 1,979 cases and 142,627 controls of European ancestry. Genotyping was carried out at deCODE Genetics. X chromosome summary statistics were only present for the Icelandic component. We used this sample to establish that SNP associations from the primary GWAS replicated *en masse* in an independent sample (see **Supplementary Note**) by showing the directions of effect of index SNPs differed from the null hypothesis of randomly oriented effects and also comparing the expected number of same direction effects with those if all associations were true, taking into account the discovery magnitude of effect, and the replication effect-estimate precision (**Supplementary Note**).

The summary statistics from deCODE were combined with those from our primary GWAS dataset using an inverse variance-weighted fixed effects model. Similarily to the discovery meta-analysis (see above) we merged overlapping LD-clumps to a total of 270 distinct genomic regions (5 on the X-chromosome) with at least one genome-wide significant signal.

### Polygenic Prediction

We estimated the cumulative contribution of SNPs to polygenic risk of schizophrenia using a series of 89 leave-one-out polygenic prediction analyses based on LD-clumping and P-value thresholding (P+T) ^11^ using PLINK^8^. For calculating polygenic scores, we included the most significant SNP for any pair of SNPs within <500kb and with LD R^2^ >0.1. We included only those with minor allele frequency >1%. We considered a range of P-value thresholds; 5×10^-8^, 1×10^-6^, 1×10^-4^, 1×10^-3^, 1×10^-2^, 5×10^-2^, 1×10^-1^, 2×10^-1^, 5×10^-1^ and 1.0. We performed logistic regression analysis within each case-control sample, to assess the relationship between case status and PRS (P+T) quantiles. The same principal components used for each GWAS were used as covariates for this analysis. Whenever the number of controls at a quantile was fewer than 5 times the number of covariates^12^, or if the higher bound for the PRS Odds Ratio (OR) became infinity, Firth’s penalised likelihood method was used to compute regression statistics, as implemented in the R package “logistf”^13^. ORs from these calculations were then meta-analysed using a fixed-effects model in the R package “metafor”^14^. To ensure stability of the estimates, meta-analysis was conservatively restricted to the 51 case-control samples which contained more than 10 individuals in the top 1% PRS, with at least one of them being a control. Analogous analyses were conducted to assess the ORs between individuals at the top and bottom quantiles. To assess the performance of PRS as a predictor of schizophrenia case status, a combined area under the receiver operating characteristic curve (AUROC) was estimated using the non-parametric meta-analysis implemented in the R package “nsROC”^15^.

### Mendelian Randomisation

We identified 58 publicly available sets of GWAS summary statistics suitable for bidirectional 2-sample Mendelian randomisation^16^. We used the IEU GWAS summary database (https://gwas.mrcieu.ac.uk/datasets/) for complex traits and diseases, with additional manual curation (SNP rsid, chromosome, position, reference and alternate alleles, effect size, standard errors, and P values) (see **Supplementary Table 6** for traits and references). We included only GWAS with over 10,000 samples, >1,000,000 SNPs. We performed QC including matching reference and alternate alleles and flipped stands to ensure all effect sizes in the GWAS studies are relative to the same allele (for strand ambiguous SNPs, if MAF ≤0.42, allele frequencies were used determine the strand, otherwise they were discarded). All GWAS were based on European ancestry samples (including schizophrenia).

To extract independent genetic instruments, we performed LD-clumping using 1000 Genomes European super-population subjects as the LD reference, with an R^2^ t hreshold of 0.001 and a window of 10Mb. We excluded the extended MHC region (chromosome 6:25-35 Mb) to avoid bias by pseudo-pleiotropy due to extensive LD in this region. We extracted 153 independent genome-wide significant SNPs for schizophrenia (EUR sample only). For each exposure-outcome pair, we used independent genome-wide significant SNPs for the exposure as instrumental variables (IV) and extracted the effects of these SNPs on the exposure and the outcome. We excluded exposure-outcome pairs with less than 10 IVs. Due to the differences in the source GWAS summary statistics, especially the imputation reference and sample sizes, IV for each exposure-outcome pair varied from 11 to 359 (**Supplementary Table 6)**.

We used several methods to estimate causal effects while detecting and correcting for horizontal pleiotropy. We used inverse variance weighted meta-analysis to incorporate causal effect estimates from multiple IVs for each exposure-outcome pair (and Maximum likelihood estimates as a supplement) ^17^ IVW meta-analysis in MR is prone to bias due to horizontal pleiotropy so we used MR-Egger regression^18^ to detect average pleotropic bias, modified Q test and Q’ test^19^ and MR-PRESSO global test^20^ to detect overall pleiotropic bias through outlier detection, and MR-PRESSO outlier test to identify specific genetic variants with horizontal pleiotropy. Furthermore, we used MR-PRESSO to correct for pleiotropic bias by removing outlier IVs in IVW meta-analysis. We adjusted MR-PRESSO results for multiple testing by Bonferroni correction with the total number of exposure-outcome pairs tested (N=116). As sensitivity analyses, we also performed MR-Egger regression to estimate causal effects while correcting for average pleiotropic bias and weighted median estimation to provide consistent causal effect estimates when at least 50% of IVs are valid. Causal effect estimates and bias detection test results are presented. Scatter plots were used to inspect the distribution of genetic effects of the SNP instruments used in each MR analysis (**Supplementary Figure 11**). We used MR-PRESSO as our primary method to assess causal effects between exposure-outcome pairs and potential pleiotropic biases. Further comments on the application of MR are provided in the **Supplementary Note**.

### Gene Set Enrichments

#### Tissue and cell types

We collected bulk RNA-seq data across 53 human tissues (GTEx v8, median across samples)^21^; from a study of 19,550 nuclei from frozen adult human post-mortem hippocampus and prefrontal cortex representing 16 different cell types^22^; from a study of ~10,000 single cells from 5 mouse brain regions (cortex, hippocampus, hypothalamus, midbrain and striatum, in addition to specific enrichments for oligodendrocytes, dopaminergic neurons, serotonergic neurons and cortical parvalbuminergic interneurons) that identified 24 cell types^23^; from a study of~500,000 single cells from the mouse nervous system (19 regions) that identified 265 cell types^24^.

Datasets were processed uniformly^25^. First, we calculated the mean expression for each gene for each type of data if these statistics were not provided by the authors. We used the precomputed median expression (transcript per million (TPM)) across individuals for the GTEx tissues (v8). For the GTEx dataset, we excluded tissues with less than 100 samples, merged tissues by organ (with the exception of brain tissues), excluded non-natural tissues (e.g. EBV-transformed lymphocytes) and testis (outlier in hierarchical clustering), resulting in 37 tissues. Genes without unique names and genes not expressed in any cell types were excluded. We scaled the expression data to 1M Unique Molecular Identifiers (UMIs) or TPM for each cell type/tissue. After scaling, we excluded non-protein coding genes, and, for mouse datasets, genes that had no expert curated 1:1 orthologs between mouse and human (Mouse Genome Informatics, The Jackson laboratory, version 11/22/2016). We then calculated a metric of gene expression specificity by dividing the expression of each gene in each cell type/tissue by the total expression of that gene in all cell types/tissue, leading to values ranging from 0 to 1 for each gene (0: meaning that the gene is not expressed in that cell type/tissue, 1 that 100% of the expression of that gene is performed in that cell type/tissue). We selected the 10% most specific genes per cell type (or tissue) with an expression level of at least 1TPM, or 1 UMI per million, for downstream analyses and used MAGMA^26^ to test whether they were enriched for genetic associations. We performed a one-sided test as we were only interested in enrichments for genetic associations (in contrast with depletions). We also applied partitioned LD score regression (LDSC) as described^27^ to the top 10% genes for each cell type for heritability enrichment. We selected the one-sided coefficient z-score p-value as a measure of the association of the cell type/tissue with schizophrenia.

#### Ontology Gene sets

Gene set analyses were performed using MAGMA v1.06^26^. Gene boundaries were retrieved from Ensembl release 92 (GRCh37) using the “biomaRt” R package ^28^ and expanded by 35 kb upstream and 10 kb downstream to include likely regulatory regions^29^. Gene-wide p-values were calculated from European and Asian summary statistics separately, and metaanalysed within the software. LD reference data files were from the European and East Asian populations of the Haplotype Reference Consortium^30^. We performed 100,000 permutations to estimate p-values corrected for multiple testing under the family-wise error rate (FWER) approach^31^. Within each collection of gene sets, we carried out stepwise conditional analyses as described ^32^, selecting the most significant set in that collection, which was then added as a covariate to a regression with all other sets. This was repeated until none of the remaining sets showed a nominally significant uncorrected p-value. Specifically, we tested the following gene sets:

i. Gene ontology: 7,083 sets extracted from the GO database (http://geneontology.org/, accession date: 04/07/2018) curated to include only annotations with experimental or phylogenetic supporting evidence. Gene sets and automated curation pipeline are provided in https://github.com/janetcharwood/pgc3-scz_wg-genesets
ii. SynGO ontology: Described 33, this collection was analysed as two subsets; “biological process” (135 gene sets) and “cellular component” (60 gene sets). We controlled for a set of 10,360 genes with detectable expression in brain tissue measured as Fragments Per Kilobase of transcript per Million mapped reads (FPKM)34 to detect synaptic signals above signals simply reflecting the property of brain expression. Gene sets were reconstructed using a “roll-up” method, in which parent categories contained all genes annotated to child categories. For conditional testing, as the regression covariate, we prioritised more specific child annotations^35^.

### Conditional Analyses

We performed stepwise conditional analyses of the 270 loci looking for independent associations. We performed association testing and meta-analysis across each locus, adding the allele dosages of the index SNP as a covariate. Where a second SNP had a conditional p-value of less than 1×10^-6^, we considered this as evidence for a second signal and repeated the process adding this as an additional covariate. We repeated this until no additional SNPs in the region achieved p<1×10^-6^. We also searched for long range dependencies. Here we tested the all pairs of independent signals for conditional independence (**Supplementary Note)**.

### Fine-mapping

We used FINEMAP^36^ to fine-map loci, excluding the xMHC region due to its complex LD structure. To simplify the causal SNP content of regions ^37^, for loci formed from nonoverlapping clumps which conditional analysis indicated contained independent signals, we fine-mapped those clumps separately. A region in chromosome 5 with long-range LD dependencies (chr5:46-50Mb) was analysed as a single locus. As fine-mapping requires data from all markers at the locus^38^ we used only included loci/clumps that attained genome-wide significance (GWS) in the discovery sample. To be conservative, we excluded 13 GWS clumps in the discovery sample that were not GWS in the replication data, although these are most likely true positives. In total, we attempted to fine-map 233 regions. Further details about the fine-mapping process are given in the **Supplementary Note**.

### Summary-data-based Mendelian Randomization (SMR) analysis, FUSION and EpiXcan

We used SMR^39^ as our primary method to identify SNPs which might mediate association with schizophrenia through effects on gene expression. The significance for SMR is set at the Bonferroni corrected threshold of 0.05/M where M is the number of genes with significant eQTLs tested for a given tissue. Significant SMR associations imply colocalization of the schizophrenia associations with eQTL. We applied the HEIDI test^39^ to filter out SMR associations (*P*_HEIDI_ < 0.01) due to linkage disequilibrium between SCZ-associated variants and eQTLs. cis-eQTL summary data were from three studies: fetal brain (N=120)^40^, adult brain (*n* = ~1,500)^41^ and blood(*n* = ~32,000)^42^. Linkage disequilibrium (LD) data required for the HEIDI test^39^ were estimated from the Health and Retirement Study (HRS)^43^ (*n* = 8,557). We included only genes with at least one *cis*-eQTL at *P*_eQTL_ < 5×10^−8^, excluding those in MHC regions due to the complexity of this region. For blood, we included only genes with eQTLs in brain. This left 7,803 genes in blood, 10,890 genes in prefrontal cortex and 754 genes in fetal brain for analysis (see **Supplementary Note** for further details).

For genomic regions where there were multiple genes showing significant SMR associations, we attempted to resolve these with conditional analysis using GCTA-COJO^44,45^. We selected the top-associated cis-eQTL for one gene (or a set of genes sharing the same *cis*-eQTL) ran a COJO analysis in the schizophrenia GWAS data and the eQTL data for each of the other genes conditioning on the selected top *cis*-eQTL. We then re-ran the SMR and HEIDI analyses using these conditional GWAS and eQTL results.

We used FUSION ^46^ and EpiXcan ^47^ as tests of robustness of the SMR results. Details are supplied in the **Supplementary Note** as are our approaches to prioritising SMR associated genes.

## Notes

### Competing Interest Statement

Aarno Palotie is a member of Astra Zenecas Genomics Advisory Board. Veikko Salomaa has consulted for Novo Nordisk and has ongoing research collaboration with Bayer Ltd (both unrelated to the present study). Michael Green is a paid consultant for AiCure, Biogen, Lundbeck, and Roche, is a member of the Scientific Board of Cadent, and has received research funds from Forum. Gregory Light has consulted to Astellas, Forum, and Neuroverse Keith Nuechterlein has research support from Janssen, Genentech, and Brain Plasticity Inc. Also has consulted to Astellas, MedinCell, Takeda, Teva, Genentech, Otsuka, Janssen, and Brain Plasticity Inc. David Cohen has reported past consultation for or the receipt of honoraria from Otsuka, Shire, Lundbeck, Roche and Janssen. Mark Daly is a founder of Maze Therapeutics. Anil K. Malhotra is a consultant to Genomind Inc, InformedDNA, and Concert Pharmaceuticals. Rodrigo Affonseca Bressan has received research grants from Janssen; has been a forum consultant for Janssen and Sanof; Roche; speaker bureau for Ache, Janssen, Sanofi and Torrent. Cristiano Noto was on the speakers' bureau and/or has acted as a consultant for Janssen and Daiichi-Sankyo in the last 12 months. Christos Pantelis has, for the last 3 years, served on an advisory board for Lundbeck and received honoraria for talks presented at educational meetings organized by Lundbeck. David A Collier is a full-time employee and stockholder of Eli Lilly and Company. Michael O'Donovan is supported by a collaborative research grant from Takeda Pharmaceuticals. Michael Owenis supported by a collaborative research grant from Takeda Pharmaceuticals. James Walters is supported by a collaborative research grant from Takeda Pharmaceuticals. Andrew Pocklington is supported by a collaborative research grant from Takeda Pharmaceuticals. Stephen R. Marder has consulted for the following companies: Roche, Sunovion, Lundbeck, Boeringer-Ingelheim, Acadia, and Merck. Srihari Gopal is a full time employee and shareholder Johnson & Johnson (AMEX: JNJ). Adam Savitz is an employee of Janssen Research & Development, LLC and own stock/stock options in the company. Qingqin Li is an employee of Janssen Research & Development, LLC and own stock/stock options in the company. Tony Kham-Thong is an employee of F.Hoffman-La Roche Anna Rautanen is an employee of F.Hoffman-La Roche Dheeraj Malhotra is an employee of F.Hoffman-La Roche Sara Paciga is an employee of Pfizer Inc.

### Author Declarations

The ethical details of each of the 90 studies are given in the supplementary "Cohort Descriptions".

